# Covid19 2020 projection report: Germany, Israel, Italy and USA. Mid-July 2020

**DOI:** 10.1101/2020.07.21.20131664

**Authors:** Isaac Meilijson

## Abstract

A notion of implied susceptible population size ISPS was introduced in the context of the SIR differential equations in Epidemiology, in a companion paper. It is the potential target population size for which the solution to the SIR equations would yield the current number of new affected cases. This notion is applied to the analysis and projection of Covid19 2020, illustrated on the data of Germany, Israel, Italy and USA.

## 1 Introduction

The SIR (Susceptibles, Infected, Removed) model introduced by Kermack & McKendrick in 1927 ([6]) for the progress of an epidemic describes the interdependence between the cumulative number *X*(*t*) of affected cases, the cumulative number *R*(*t*) of removed cases (dead or recovered) and the ensuing current number of infected cases *I*(*t*) = *X*(*t*) − *R*(*t*). The version of this model to be applied in the current report conforms with the usual assumption *dR*(*t*) = *γI*(*t*)*dt* that the number of new removed cases constitutes a fixed proportion of the currently infected cases, and the usual assumption that the number of new affected cases is proportional to the product of the number *K* − *X*(*t*) of susceptible cases and an increasing function of the number of currently infected cases. However, *K* is fitted from data instead of letting it be the total population size, and the second factor (commonly modelled as the identity function) is taken (after Grenfell, Bjørnstad & Filkenstädt ([4])) to be a fractional power *I*(*t*)^*α*^ (0 ≤ *α* < 1) of the number of currently infected cases. In other words, letting 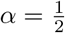,

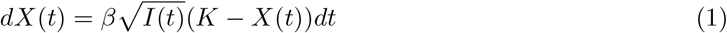

The companion publications by Alon and/or the author ([1], [7]) to the current report describe methods to estimate the parameters *α, β, γ* and *K*. It also describes a regression pre-processing procedure that modifies minimally the infected and removed components of the empirical data (*X*_1_, *I*_1_, *R*_1_),

(*X*_2_, *I*_2_, *R*_2_), …, (*X*_*n*_, *I*_*n*_, *R*_*n*_), so that the proportionality of new removals and currently infected cases will better hold. This procedure ignores equation (1).

The data analyzed in the current report has been submitted to this pre-processing procedure, and maximum likelihood estimation based on the SIR differential equations has led to the estimation of the four parameters in a period considered stationary, days 60 to 120 in the monitoring of Covid19 2020. The working paradigm is that the three parameters *α, β, γ* hold more generally. Under this assumption, equation (1) is rewritten as

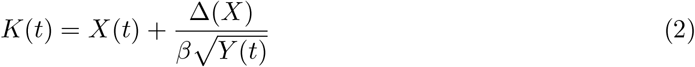

to be interpreted as the *implied susceptible population size* ISPS. Letting Δ(*X*) be a smooth version of *X*(*t* + 1) − *X*(*t*), K(t) is evaluated daily to provide an idea of the current target population of the epidemic. Due to the persistent missing data on weekends, the smooth version is taken throughout to be a moving average on the week around the date in question.

### A word on the parameter *α*

Imagine a small-world network topology composed of sparsely connected islands that are internally densely connected. Once an island is affected, contagion will apply locally. Much as in the Harris contact process ([5]), if the affected cases are considered a growing ball that transmits the disease by its perimeter, 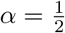 models the intra-island spread force. The “ball” could have some fractal dimension rather than a solid area. [1] presents an argument that on infinite populations, *α* = 1 yields exponential growth of *X*, while *α* < 1 gives rise to linear growth of *X*, with *I*(*t*) increasing asymptotically to oscillate around the finite constant 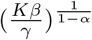. (Formula (1) explains why should the proper parameter be *Kβ* rather than *β*, for a large population).

The average daily number of new affected and removed cases would then be 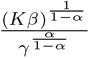. These levels, to be reported for every country under study, are so high that it is reasonable to infer that only a small fraction of the population is susceptible.

Without claiming to delve deeply into Epidemiology, it is suggested that isolation measures achieve stationarity by cutting contagion across islands. Relaxing isolation measures may have the effect of allowing for more contagion between islands, and this is what ISPS is intended to reflect.

In each of the four countries that illustrate the analysis in the current report, the value 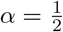 is solidly in the confidence interval for *α*, as dictated by a profile likelihood function. This value will be applied throughout.

Since (the ideally constant) ISPS function is viewed as a stochastic process, it is convenient to monitor its logarithmic daily rate of growth function “Delta Log K” 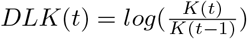. Under stationary behavior, this function should in principle be zero and in practice it should oscillate around zero. It should be stressed that a period of (positive) constancy of DLK signifies exponential growth of the target population.

This is apparent in Figures 3 and 6, that also illustrate a scenario beyond the monitoring period, under which *DLK* stays constant for 10 days and then decays exponentially at daily rate 2.5%. As this scenario calls for a reversal of the increasing trend of *DLK* in Israel and the USA, it is intended as an optimistic scenario, however grim its projected consequences may appear.

Israel went through a Covid19 first wave that seemed to saturate, but relaxation of measures brought the current second wave. USA is (perhaps wrongly) treated as one unit, and as such never quite stabilized. Unlike Israel and USA, the two countries Germany and Italy (similarly Switzerland) seem to have maintained beyond the training period a combination of official isolation measures and personal responsibility that has prevented so far a return to the pandemic core.

The purpose of this report is to suggest possible uses of ISPS as a measure of growth and perhaps as a driving wheel of epidemic management, without taking sides on discussions about management. With this said, under the strong evidence that *α* < 1, the author rejects the postulate of exponential growth in homogeneous sub-populations.

Evaluation of ISPS and DKL will be illustrated on four countries, projecting into the near future the solution to the SIR equations, with *β, γ* and 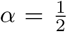 as fitted in the training period, and ISPS derived from the above *DLK* scenario. Figures to be presented show *ISPS* and *DLK* as well as *X, Y, R*. The *X, Y, R* data for the four countries are tabulated in detail.

### Robustness

The purpose of converting true empirical data to an implied index, ISPS in this report and implied volatility in Finance, is to reduce the effect of the model used, by adjusting to it. Projections via the scenario described above will be performed by letting *α* in (1) and (2) take all values in {0, 0.2, 0.4, 0.6, 0.8, 1}. For each such *α*, the two parameters *β* (highly changing) and *γ* (quite insensitive to *α*) will be estimated on the training period data, keeping *K* as estimated for 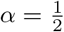. Figures 13, 14, 15 display (*X, I, R*), *DLK* and *ISPS* in Israel, for the various values of *α*.

## 2 Scenario analysis: Covid 19 in Israel

Estimation in the training period yielded *β* = 9.9636 10^−4^ and *γ* = 0.0382. That is, an infected person is removed (dead or recovered) after 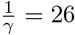 days on average. If the population of susceptible cases was infinite, there would be constantly 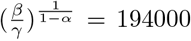 infected cases and the number of daily new affected and removed cases would be 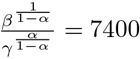.

Figure 2 shows that during the (presumably stationary) training period from day 60 to day 120, DLK oscillates around zero, progressively stable. It is essentially zero between days 105 and 120, after which isolation measures were lifted. *DLK* sharply increases and “stabilizes” around 0.015 until day 140 and then experiences a new height around 0.035 until the end of the monitoring period, day 176 (July 16, 2020). This figure also illustrates the scenario for extending *DLK* into the future - constant for 10 days followed by exponential decay at daily rate 2.5%. Figure 3 displays the corresponding function *ISPS*.

**Figure 1:**
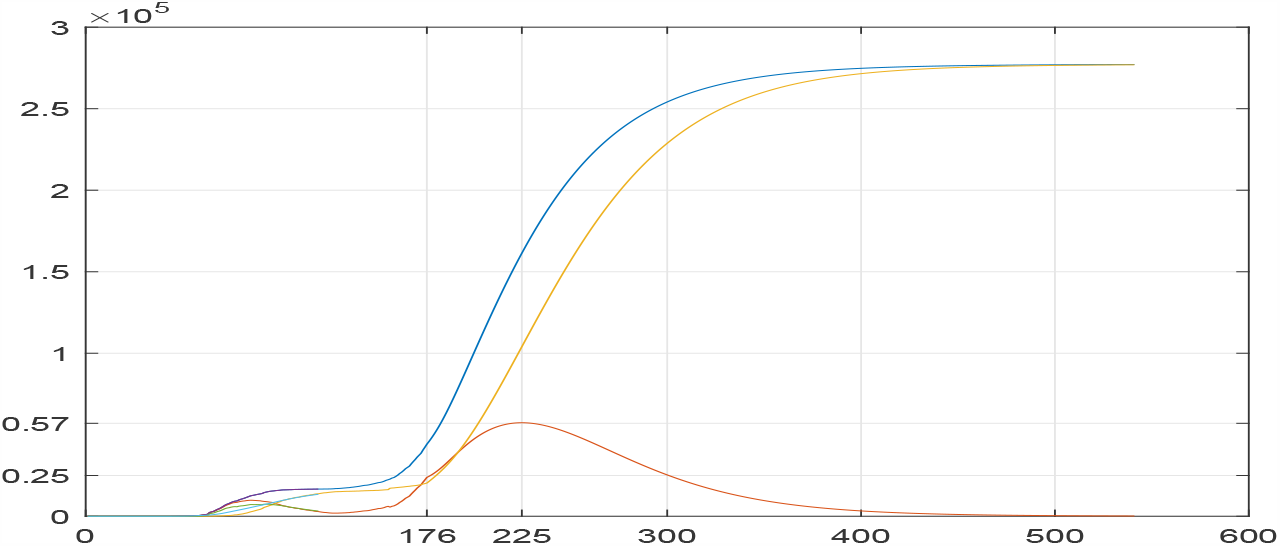
ISRAEL: Affected, currently infected, removed cases in the training (first 60 days) and observation periods (until day 176), and beyond. Data, SIR solution and scenario projection.

**Figure 2:**
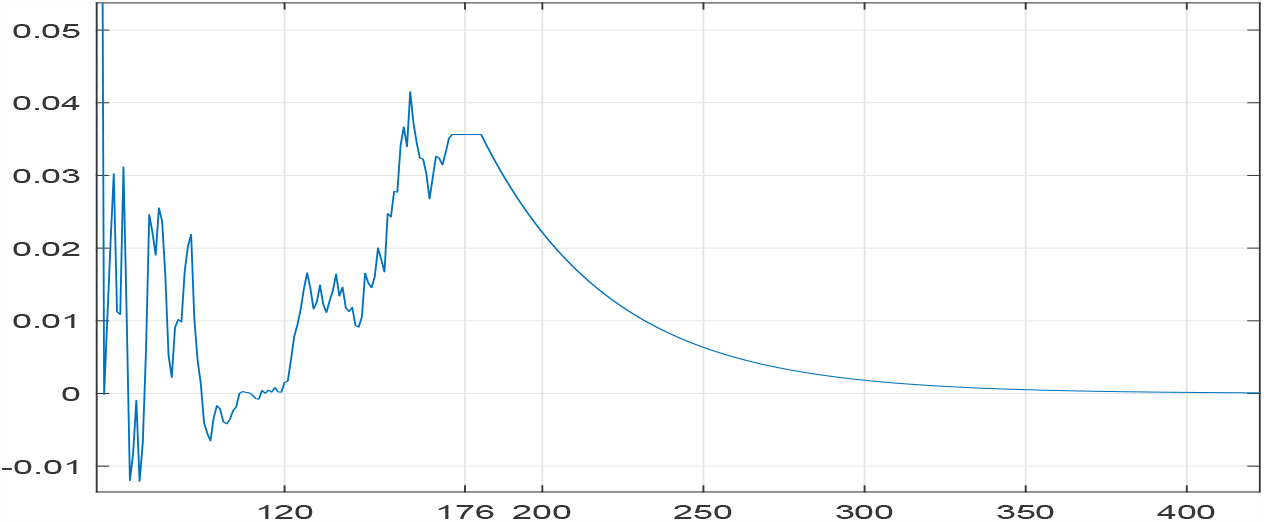
ISRAEL: Incremental logarithm *DLK* of ISPS *K*, data and scenario projection.

**Figure 3:**
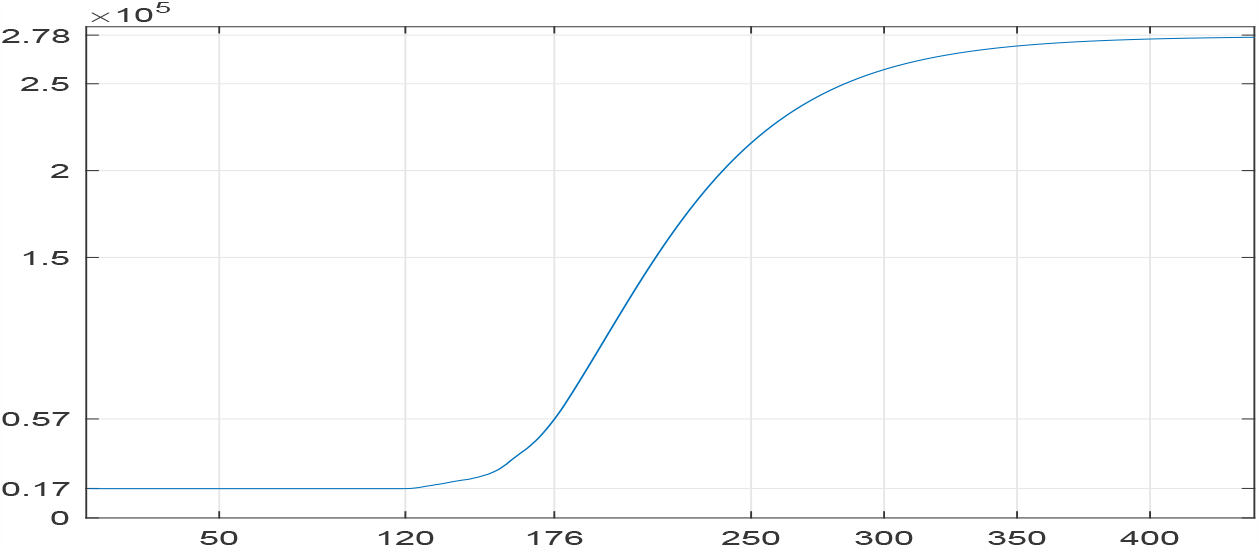
ISRAEL: ISPS *K*, data and scenario projection.

Figure 1 shows that on July 16, the cumulative number of affected cases was 44000 and the number of currently infected cases was 25000. Under the scenario above, the number of infected cases will increase to a maximum (the basic reproduction number *R*_0_ crosses from above to below 1) of 57000 on September 3rd, by which time the cumulative number of affected cases would be 161000. Under the new stationary period reflected by the scenario, the maximal number of affected cases would be 277000.

No other scenarios are displayed in figures. However, it can be reported that a delay of one day in keeping *DLK* constant in Israel before starting the exponential decay will see the maximal number of affected cases increase from 277000 by over 10000, and every further delay by one day before starting the exponential decay will see the incremental maximal number of affected cases increase by 3.56% a day. If only 1% of the removed cases die, this means that every day of delay in starting the exponential decay will bring over 100 dead cases, 25% of the current total number of dead cases 400.

## 3 Scenario analysis: Covid 19 in USA

Estimation in the training period yielded *β* = 2.839 10^−5^ and *γ* = 0.0117. That is, an infected person is removed (dead or recovered) after 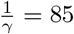 days on average. If the population of susceptible cases was infinite, there would be constantly 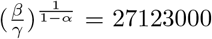 infected cases and the number of daily new affected and removed cases would be 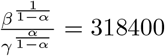.

Figure 5 shows that in USA *DLK* never quite stabilizes but tends to show mean reversion towards zero until day 110, then oscillates around 0.005 until day 140 and sharply increases to its new metastable height 0.02. This figure also illustrates the scenario for extending *DLK* into the future - constant for 10 days followed by exponential decay at daily rate 2.5%. Figure 6 displays the corresponding function *ISPS*.

**Figure 4:**
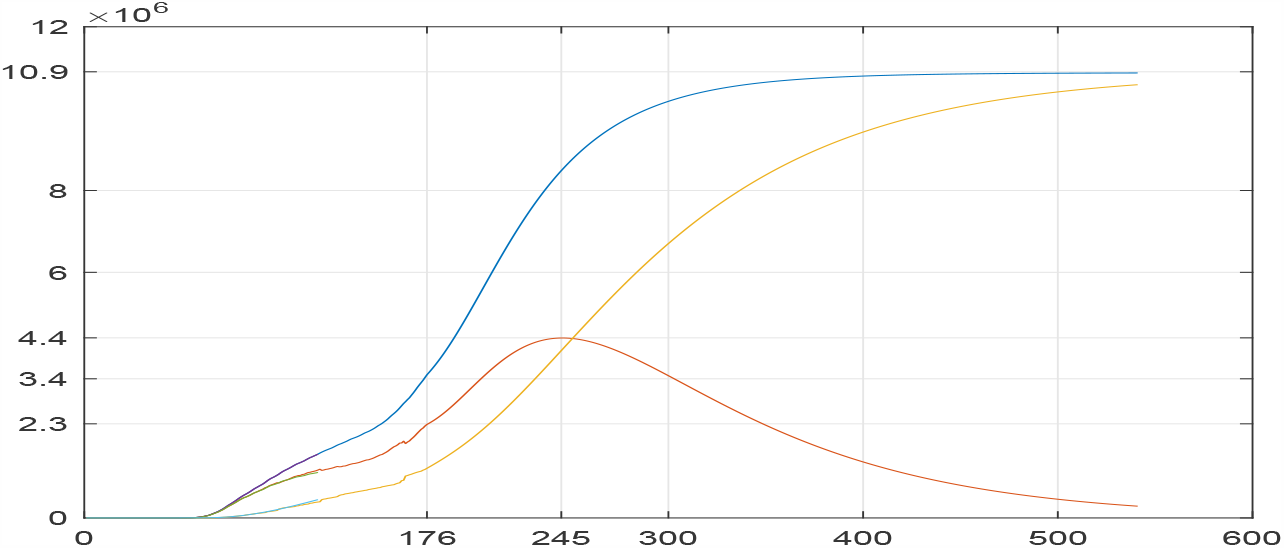
USA: Affected, currently infected, removed cases in the training (first 60 days) and observation periods (to day 176), and beyond. Data, SIR solution and scenario projection.

**Figure 5:**
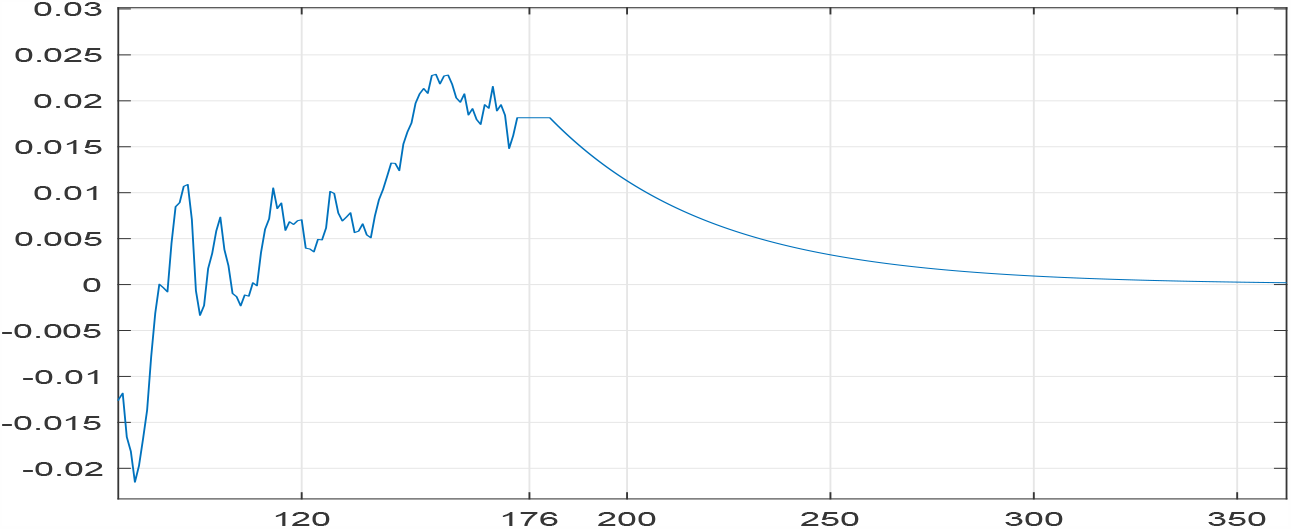
USA: Incremental logarithm *DLK* of ISPS *K*, data and scenario projection.

**Figure 6:**
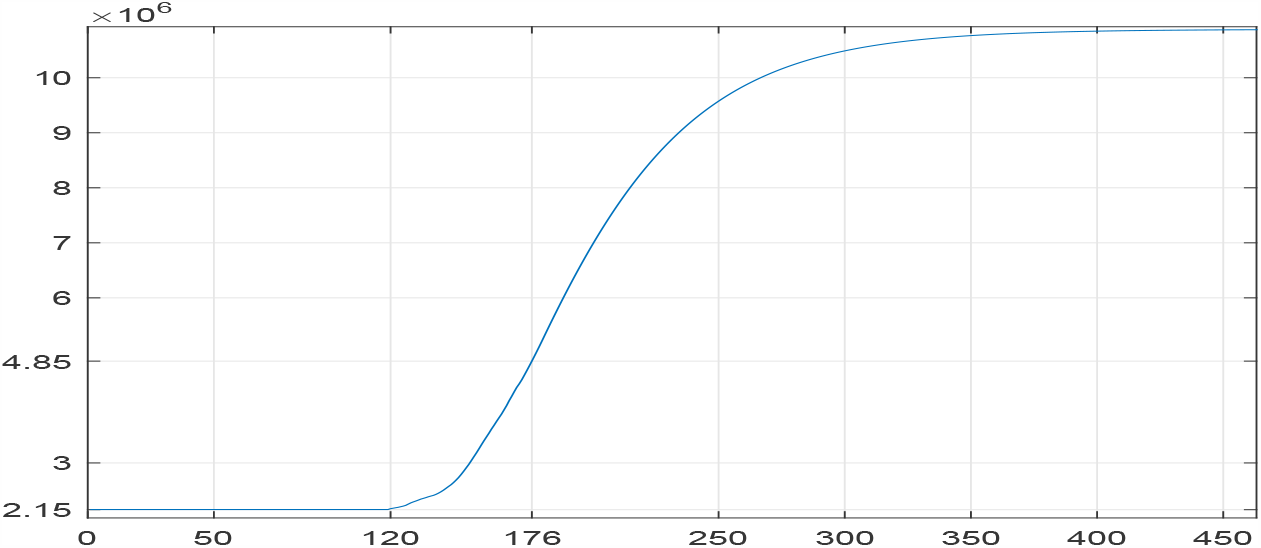
USA: ISPS *K*, data and scenario projection.

**Figure 7:**
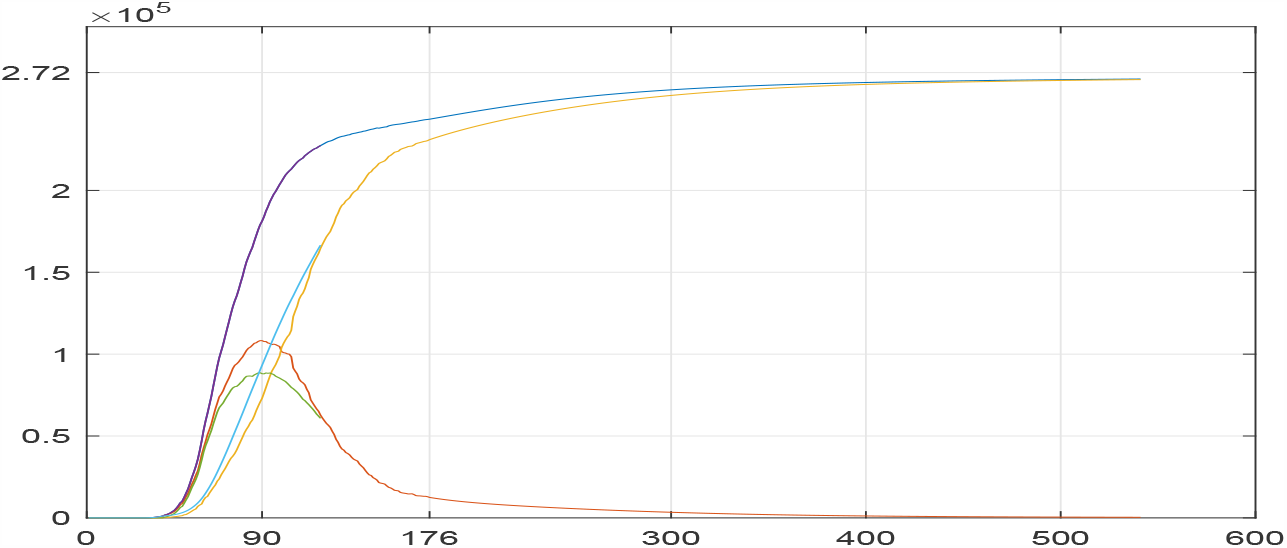
ITALY: Affected, currently infected, removed cases in the training (first 60 days) and observation periods (to day 176), and beyond. Data, SIR solution and scenario projection.

Figure 4 shows that on July 16, the cumulative number of affected cases was 3.4*M* and the number of infected cases was 2.3*M*. Under the scenario above, the number of infected cases will increase to a maximum (the basic reproduction number *R*_0_ crosses from above to below 1) of 4.4*M* on September 23rd, by which time the cumulative number of affected cases would be 8.4*M*. Under the new stationary period reflected by the scenario, the maximal number of affected cases would be 10.9*M*.

## 4 Scenario analysis: Covid 19 in Italy

Estimation in the training period yielded *β* = 9.9636 10^−4^ and *γ* = 0.0321. That is, an infected person is removed (dead or recovered) after 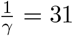 days on average. If the population of susceptible cases was infinite, there would be constantly 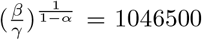 infected cases and the number of daily new affected and removed cases would be 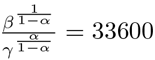.

Figure 8 shows that in Italy *DLK* stably oscillates around zero in the entire observation period. The scenario for extending *DLK* into the future - constant for 10 days followed by exponential decay at daily rate 2.5%, may be a random effect. ISPS was estimated as 250000 during the training period and never exceeded 270000.

**Figure 8:**
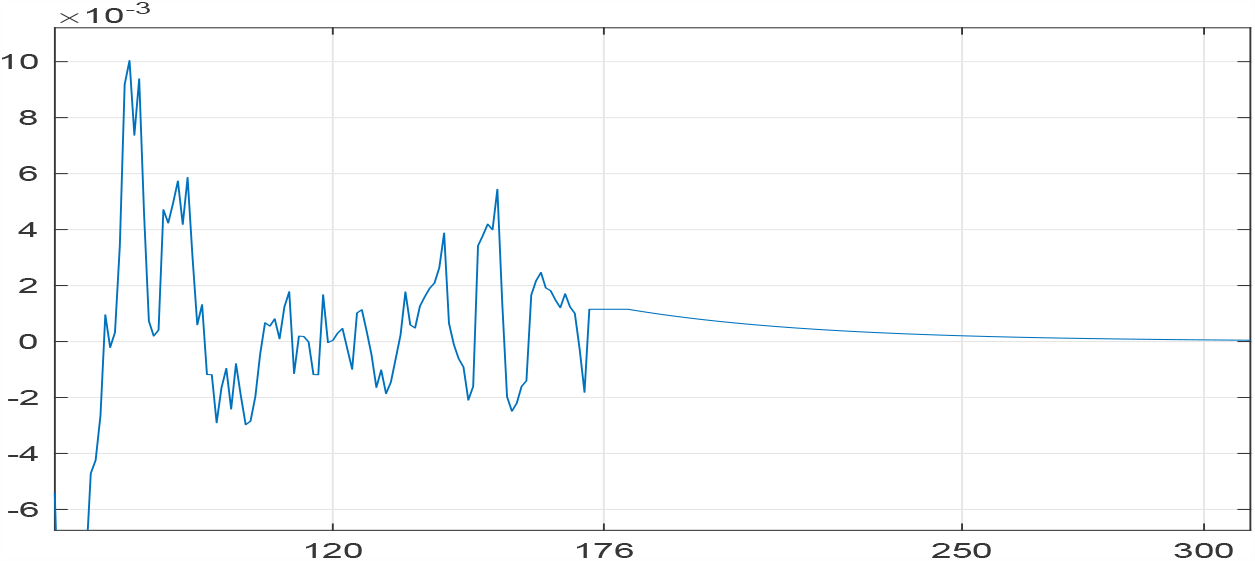
ITALY: Incremental logarithm *DLK* of ISPS *K*, data and scenario projection.

**Figure 9:**
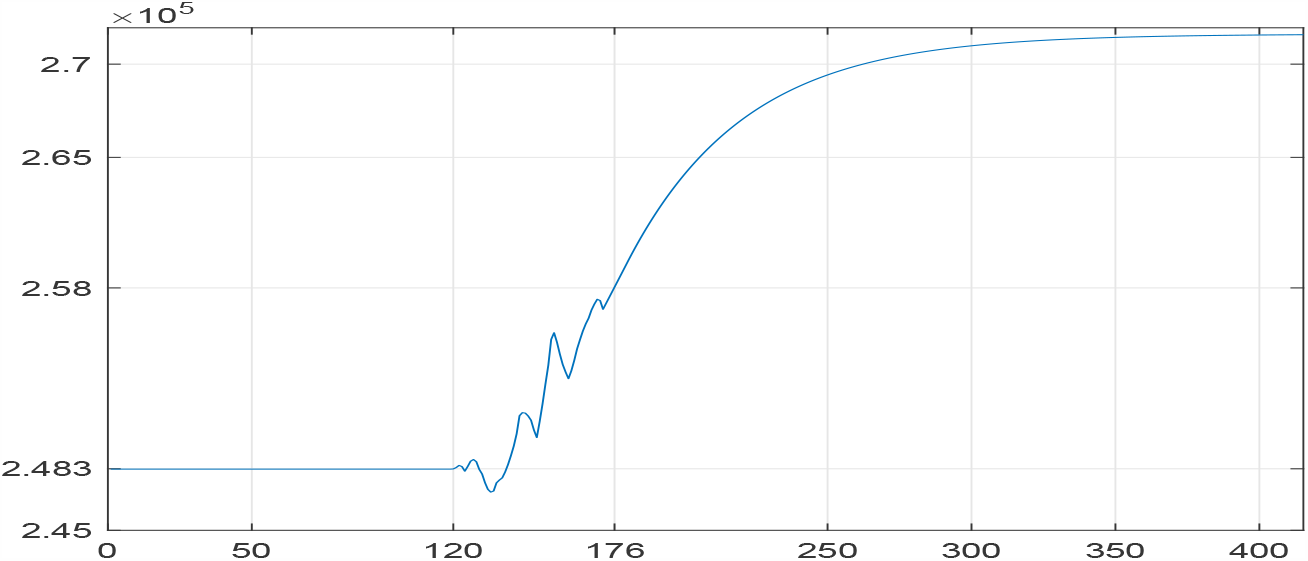
ITALY: ISPS *K*, data and scenario projection.

**Figure 10:**
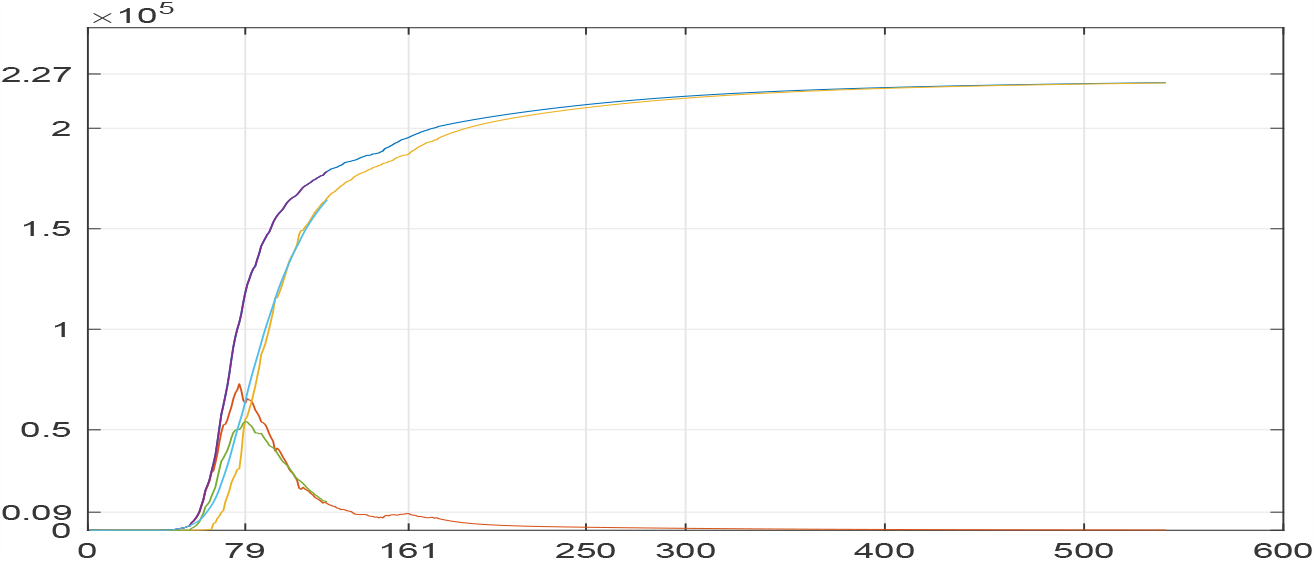
GERMANY: Affected, currently infected, removed cases in the training (first 60 days) and observation periods (to day 176), and beyond. Data, SIR solution and scenario projection.

The maximal number of infected cases was around 100000, reached on day 90, April 21st. Affected cases are accurately fitted, but there is some disparity between the way empirical data and SIR solutions split the affected cases into infected and removed.

## 5 Scenario analysis: Covid 19 in Germany

Estimation in the training period yielded *β* = 9.9636 10^−4^ and *γ* = 0.0774. That is, an infected person is removed (dead or recovered) after 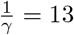 days on average. If the population of susceptible cases was infinite, there would be constantly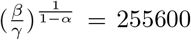 infected cases and the number of daily new affected and removed cases would be 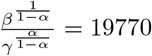.

Figure 11 shows that in Germany *DLK* stably oscillates around zero in the entire observation period. Germany experienced a mini second wave with a local maximum of infected cases at day 161, that shows itself in a discontinuous jump in ISPS. Except for this, the scenario for extending *DLK* into the future - constant for 10 days followed by exponential decay at daily rate 2.5%, may be a random effect as in Italy. ISPS was estimated as 200000 during the training period, increased sharply to 215000 and then smoothly to 22700.

**Figure 11:**
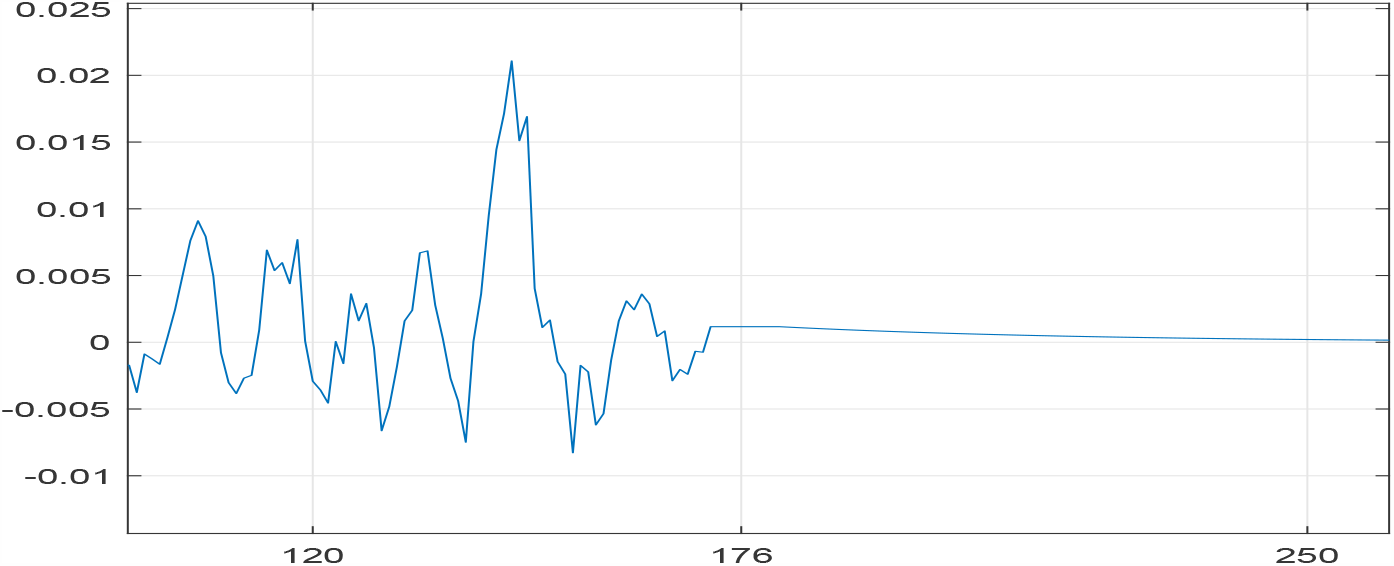
GERMANY: Incremental logarithm *DLK* of ISPS *K*, data and scenario projection.

**Figure 12:**
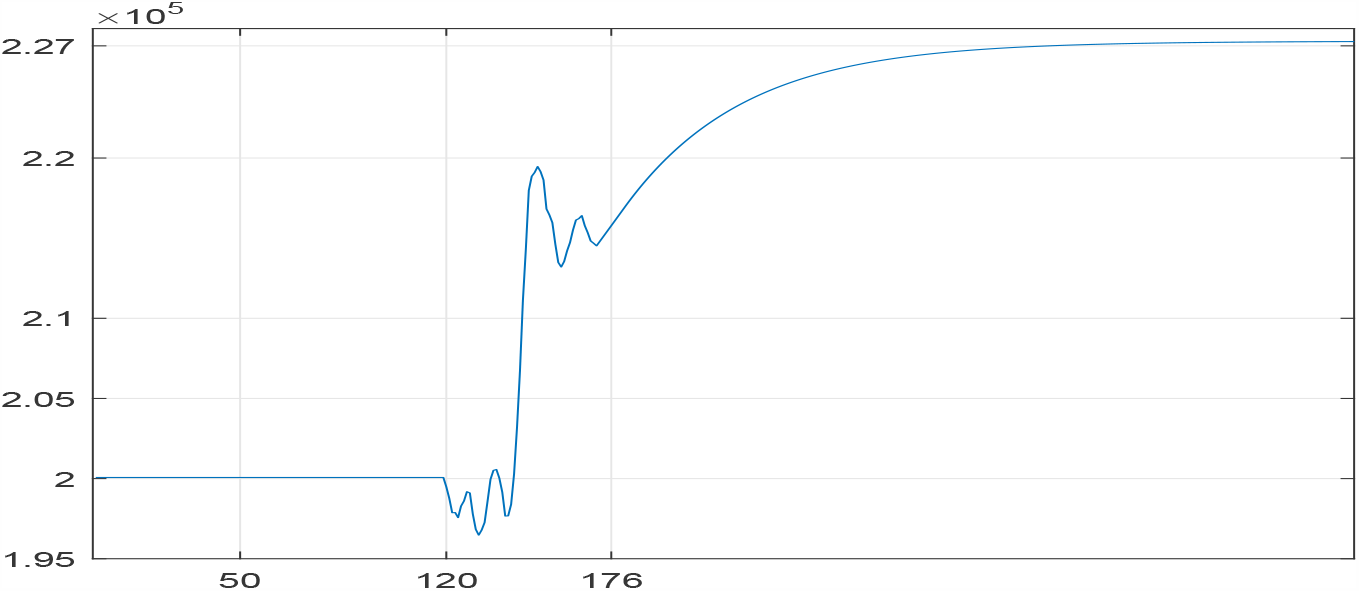
GERMANY: ISPS *K*, data and scenario projection.

The maximal number of infected cases was around 50000, reached on day 79, April 10. Affected cases are accurately fitted, but, as in Italy, there is some disparity between the way empirical data and SIR solutions split the affected cases into infected and removed.

## 6 Robustness with respect to *α*

Figures 13, 14, 15 display (*X, I, R*), *DLK* and *ISPS* in Israel, for *α* covering the entire range [0, 1] and *K* frozen at the value estimated under 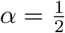. The values of *β* and *γ* are best fitted for each given *α*. The parameter *γ* increases slowly from 0.0368 to 0.0388 as *α* increases from 0 to 0.6, and then to 0.0408 and 0.0444 at *α* = 0.8 and 1.0. As a curiosity, the parameter *β* is practically identical to the function 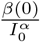 where the solution for *α* = 0 is *β*(0) = 1207 and the other constant is *I*_0_ = 4927.

**Figure 13:**
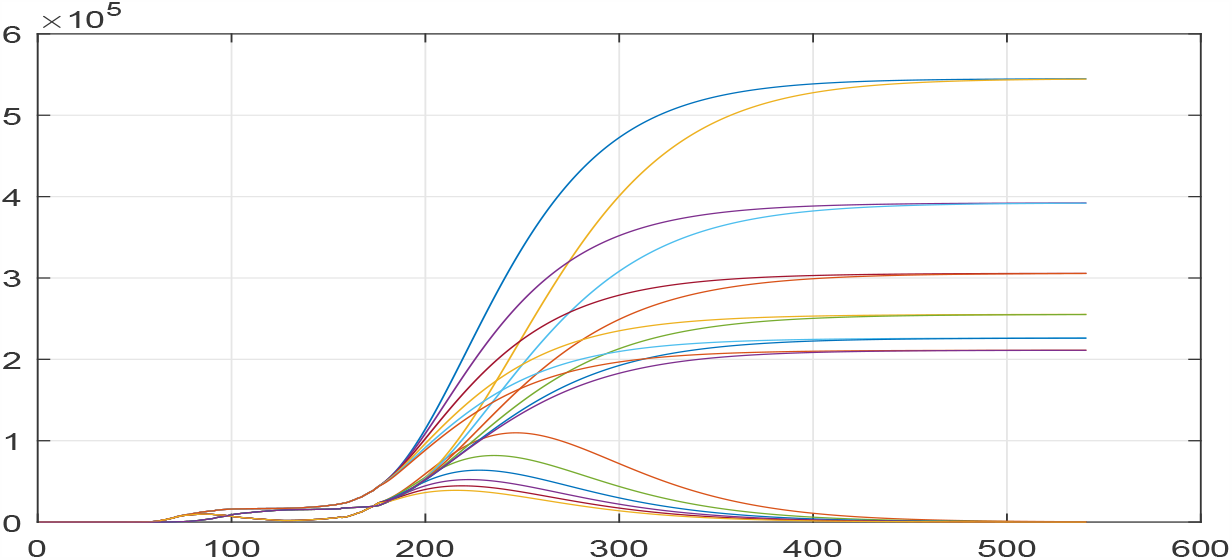
ISRAEL: Affected, currently infected, removed cases under different *α*. Functions decrease from *α* = 0 to *α* = 1, and are similar to each other for *α ≥* 0.4.

**Figure 14:**
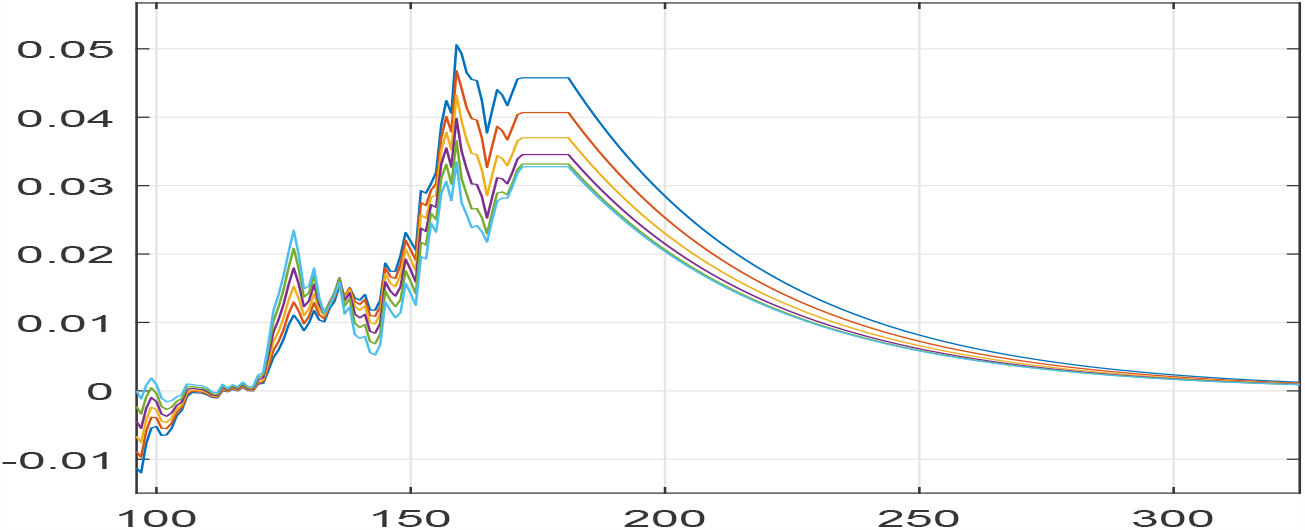
ISRAEL: Incremental logarithm *DLK* of ISPS *K*, under different *α*.

**Figure 15:**
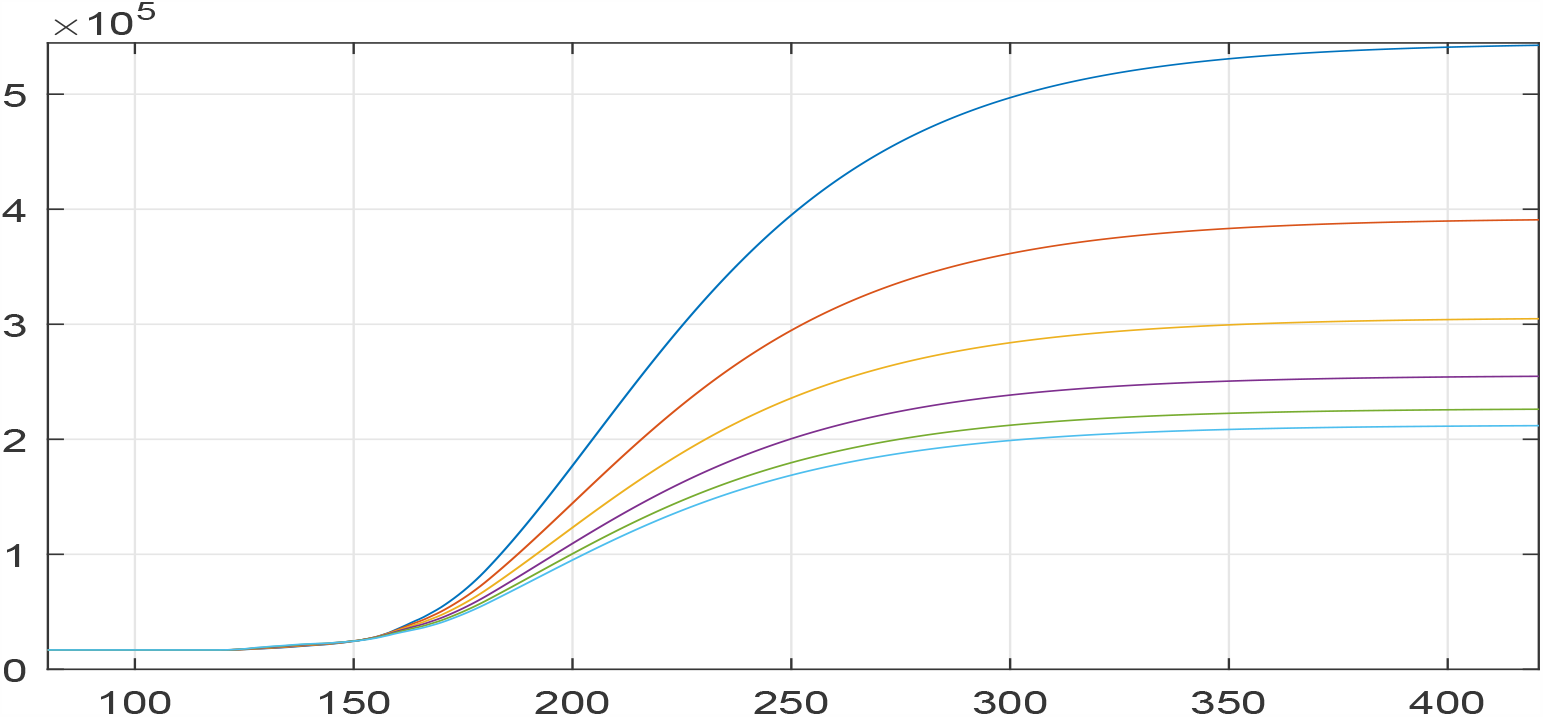
ISRAEL: ISPS *K*, under different *α*.

So, if equation (1) had been written in terms of 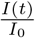 in place of *I*(*t*), the parameter *β* would have been the same for all *α*. Material left for further thought.

It is apparent that the two top functions in Figure 13, corresponding to *α* = 0 (affected cases increase proportionally to the susceptible cases only) and to *α* = 0.2 are off, but all others give similar assessments to all three processes *X, I* and *R*.

## Data Availability

Johns Hopkins data

## 7 Tables

**Table 1:**

|  |  | GERMANY |  |  |  | GERMANY |  |  |
| --- | --- | --- | --- | --- | --- | --- | --- | --- |
| Date | Affected | Infected | Removed |  | Date | Affected | Infected | Removed |
| 30-Jun-20 | 195042 | 8296 | 186746 |  | 14-Aug-20 | 206078 | 2563 | 203516 |
| 01-Jul-20 | 195418 | 8328 | 187090 |  | 15-Aug-20 | 206229 | 2517 | 203712 |
| 02-Jul-20 | 195893 | 7798 | 188095 |  | 16-Aug-20 | 206379 | 2474 | 203905 |
| 03-Jul-20 | 196370 | 7564 | 188806 |  | 17-Aug-20 | 206527 | 2432 | 204095 |
| 04-Jul-20 | 196780 | 7470 | 189310 |  | 18-Aug-20 | 206674 | 2392 | 204281 |
| 05-Jul-20 | 197198 | 7178 | 190020 |  | 19-Aug-20 | 206819 | 2354 | 204465 |
| 06-Jul-20 | 197523 | 6781 | 190742 |  | 20-Aug-20 | 206963 | 2317 | 204646 |
| 07-Jul-20 | 198064 | 6882 | 191182 |  | 21-Aug-20 | 207106 | 2282 | 204824 |
| 08-Jul-20 | 198343 | 6650 | 191693 |  | 22-Aug-20 | 207247 | 2248 | 204999 |
| 09-Jul-20 | 198699 | 6500 | 192199 |  | 23-Aug-20 | 207388 | 2216 | 205172 |
| 10-Jul-20 | 199001 | 6216 | 192785 |  | 24-Aug-20 | 207527 | 2185 | 205342 |
| 11-Jul-20 | 199332 | 6241 | 193091 |  | 25-Aug-20 | 207665 | 2155 | 205510 |
| 12-Jul-20 | 199709 | 6373 | 193336 |  | 26-Aug-20 | 207801 | 2126 | 205675 |
| 13-Jul-20 | 199919 | 6434 | 193485 |  | 27-Aug-20 | 207937 | 2098 | 205839 |
| 14-Jul-20 | 200180 | 6006 | 194174 |  | 28-Aug-20 | 208071 | 2072 | 206000 |
| 15-Jul-20 | 200456 | 6278 | 194178 |  | 29-Aug-20 | 208205 | 2046 | 206159 |
| 16-Jul-20 | 200890 | 5810 | 195080 |  | 30-Aug-20 | 208337 | 2021 | 206317 |
| 17-Jul-20 | 201102 | 5586 | 195516 |  | 31-Aug-20 | 208468 | 1996 | 206472 |
| 18-Jul-20 | 201313 | 5373 | 195940 |  | 01-Sep-20 | 208598 | 1973 | 206625 |
| 19-Jul-20 | 201521 | 5173 | 196348 |  | 02-Sep-20 | 208727 | 1950 | 206777 |
| 20-Jul-20 | 201725 | 4984 | 196741 |  | 03-Sep-20 | 208855 | 1928 | 206927 |
| 21-Jul-20 | 201926 | 4806 | 197120 |  | 04-Sep-20 | 208982 | 1906 | 207075 |
| 22-Jul-20 | 202125 | 4639 | 197485 |  | 05-Sep-20 | 209108 | 1886 | 207222 |
| 23-Jul-20 | 202320 | 4482 | 197838 |  | 06-Sep-20 | 209232 | 1865 | 207367 |
| 24-Jul-20 | 202513 | 4334 | 198179 |  | 07-Sep-20 | 209356 | 1846 | 207511 |
| 25-Jul-20 | 202704 | 4195 | 198509 |  | 08-Sep-20 | 209479 | 1826 | 207653 |
| 26-Jul-20 | 202892 | 4064 | 198828 |  | 09-Sep-20 | 209601 | 1807 | 207793 |
| 27-Jul-20 | 203078 | 3940 | 199138 |  | 10-Sep-20 | 209722 | 1789 | 207932 |
| 28-Jul-20 | 203261 | 3823 | 199438 |  | 11-Sep-20 | 209841 | 1771 | 208070 |
| 29-Jul-20 | 203442 | 3712 | 199730 |  | 12-Sep-20 | 209960 | 1754 | 208206 |
| 30-Jul-20 | 203621 | 3608 | 200013 |  | 13-Sep-20 | 210078 | 1737 | 208341 |
| 31-Jul-20 | 203798 | 3510 | 200288 |  | 14-Sep-20 | 210195 | 1720 | 208475 |
| 01-Aug-20 | 203972 | 3416 | 200556 |  | 15-Sep-20 | 210311 | 1703 | 208608 |
| 02-Aug-20 | 204145 | 3328 | 200817 |  | 16-Sep-20 | 210426 | 1687 | 208739 |
| 03-Aug-20 | 204316 | 3245 | 201071 |  | 17-Sep-20 | 210540 | 1671 | 208869 |
| 04-Aug-20 | 204485 | 3166 | 201319 |  | 18-Sep-20 | 210653 | 1656 | 208997 |
| 05-Aug-20 | 204652 | 3091 | 201561 |  | 19-Sep-20 | 210765 | 1640 | 209125 |
| 06-Aug-20 | 204817 | 3019 | 201797 |  | 20-Sep-20 | 210876 | 1625 | 209251 |
| 07-Aug-20 | 204980 | 2952 | 202028 |  | 21-Sep-20 | 210987 | 1610 | 209376 |
| 08-Aug-20 | 205142 | 2888 | 202254 |  | 22-Sep-20 | 211096 | 1596 | 209500 |
| 09-Aug-20 | 205302 | 2827 | 202475 |  | 23-Sep-20 | 211204 | 1581 | 209623 |
| 10-Aug-20 | 205460 | 2769 | 202692 |  | 24-Sep-20 | 211312 | 1567 | 209745 |
| 11-Aug-20 | 205617 | 2713 | 202904 |  | 25-Sep-20 | 211419 | 1553 | 209866 |
| 12-Aug-20 | 205772 | 2661 | 203112 |  | 26-Sep-20 | 211525 | 1539 | 209985 |
| 13-Aug-20 | 205926 | 2611 | 203315 |  | 27-Sep-20 | 211629 | 1526 | 210104 |
| 14-Aug-20 | 206078 | 2563 | 203516 |  | 28-Sep-20 | 211733 | 1512 | 210221 |

|  |  | ISRAEL |  |  |  | ISRAEL |  |  |
| --- | --- | --- | --- | --- | --- | --- | --- | --- |
| Date | Affected | Infected | Removed |  | Date | Affected | Infected | Removed |
| 30-Jun-20 | 24441 | 6904 | 17537 |  | 14-Aug-20 | 112722 | 50975 | 61747 |
| 01-Jul-20 | 25244 | 7583 | 17661 |  | 15-Aug-20 | 115342 | 51636 | 63706 |
| 02-Jul-20 | 26257 | 8483 | 17774 |  | 16-Aug-20 | 117949 | 52258 | 65690 |
| 03-Jul-20 | 27047 | 9176 | 17871 |  | 17-Aug-20 | 120540 | 52842 | 67698 |
| 04-Jul-20 | 28055 | 10060 | 17995 |  | 18-Aug-20 | 123115 | 53389 | 69726 |
| 05-Jul-20 | 29170 | 11024 | 18146 |  | 19-Aug-20 | 125672 | 53898 | 71775 |
| 06-Jul-20 | 29958 | 11677 | 18281 |  | 20-Aug-20 | 128211 | 54369 | 73842 |
| 07-Jul-20 | 30749 | 12359 | 18390 |  | 21-Aug-20 | 130731 | 54804 | 75927 |
| 08-Jul-20 | 32222 | 13653 | 18569 |  | 22-Aug-20 | 133230 | 55202 | 78028 |
| 09-Jul-20 | 33557 | 14875 | 18682 |  | 23-Aug-20 | 135708 | 55565 | 80143 |
| 10-Jul-20 | 34825 | 16025 | 18800 |  | 24-Aug-20 | 138164 | 55892 | 82271 |
| 11-Jul-20 | 36266 | 17302 | 18964 |  | 25-Aug-20 | 140597 | 56185 | 84412 |
| 12-Jul-20 | 37464 | 18296 | 19168 |  | 26-Aug-20 | 143007 | 56444 | 86562 |
| 13-Jul-20 | 38670 | 19300 | 19370 |  | 27-Aug-20 | 145393 | 56670 | 88722 |
| 14-Jul-20 | 40632 | 20872 | 19760 |  | 28-Aug-20 | 147754 | 56863 | 90890 |
| 15-Jul-20 | 42360 | 22324 | 20036 |  | 29-Aug-20 | 150089 | 57024 | 93065 |
| 16-Jul-20 | 44188 | 23823 | 20365 |  | 30-Aug-20 | 152400 | 57154 | 95245 |
| 17-Jul-20 | 45704 | 24427 | 21277 |  | 31-Aug-20 | 154684 | 57254 | 97430 |
| 18-Jul-20 | 47322 | 25099 | 22223 |  | 01-Sep-20 | 156941 | 57323 | 99618 |
| 19-Jul-20 | 49026 | 25831 | 23195 |  | 02-Sep-20 | 159172 | 57364 | 101808 |
| 20-Jul-20 | 50818 | 26622 | 24197 |  | 03-Sep-20 | 161375 | 57376 | 103998 |
| 21-Jul-20 | 52700 | 27470 | 25229 |  | 04-Sep-20 | 163550 | 57361 | 106189 |
| 22-Jul-20 | 54672 | 28376 | 26296 |  | 05-Sep-20 | 165698 | 57319 | 108379 |
| 23-Jul-20 | 56738 | 29340 | 27398 |  | 06-Sep-20 | 167818 | 57251 | 110567 |
| 24-Jul-20 | 58889 | 30351 | 28538 |  | 07-Sep-20 | 169909 | 57158 | 112751 |
| 25-Jul-20 | 61118 | 31401 | 29717 |  | 08-Sep-20 | 171972 | 57040 | 114932 |
| 26-Jul-20 | 63416 | 32479 | 30937 |  | 09-Sep-20 | 174006 | 56899 | 117107 |
| 27-Jul-20 | 65778 | 33579 | 32198 |  | 10-Sep-20 | 176012 | 56735 | 119277 |
| 28-Jul-20 | 68195 | 34693 | 33502 |  | 11-Sep-20 | 177988 | 56549 | 121440 |
| 29-Jul-20 | 70661 | 35812 | 34849 |  | 12-Sep-20 | 179936 | 56341 | 123595 |
| 30-Jul-20 | 73170 | 36932 | 36238 |  | 13-Sep-20 | 181855 | 56113 | 125742 |
| 31-Jul-20 | 75715 | 38045 | 37670 |  | 14-Sep-20 | 183744 | 55864 | 127880 |
| 01-Aug-20 | 78291 | 39147 | 39144 |  | 15-Sep-20 | 185605 | 55597 | 130008 |
| 02-Aug-20 | 80893 | 40233 | 40660 |  | 16-Sep-20 | 187437 | 55312 | 132126 |
| 03-Aug-20 | 83516 | 41299 | 42217 |  | 17-Sep-20 | 189240 | 55008 | 134232 |
| 04-Aug-20 | 86155 | 42341 | 43814 |  | 18-Sep-20 | 191015 | 54688 | 136326 |
| 05-Aug-20 | 88807 | 43356 | 45451 |  | 19-Sep-20 | 192760 | 54352 | 138408 |
| 06-Aug-20 | 91467 | 44342 | 47126 |  | 20-Sep-20 | 194478 | 54001 | 140477 |
| 07-Aug-20 | 94133 | 45295 | 48838 |  | 21-Sep-20 | 196166 | 53634 | 142532 |
| 08-Aug-20 | 96801 | 46216 | 50585 |  | 22-Sep-20 | 197827 | 53254 | 144573 |
| 09-Aug-20 | 99469 | 47102 | 52367 |  | 23-Sep-20 | 199459 | 52861 | 146598 |
| 10-Aug-20 | 102134 | 47951 | 54183 |  | 24-Sep-20 | 201064 | 52455 | 148609 |
| 11-Aug-20 | 104794 | 48764 | 56030 |  | 25-Sep-20 | 202640 | 52036 | 150604 |
| 12-Aug-20 | 107446 | 49539 | 57907 |  | 26-Sep-20 | 204190 | 51607 | 152583 |
| 13-Aug-20 | 110089 | 50276 | 59813 |  | 27-Sep-20 | 205712 | 51167 | 154545 |
| 14-Aug-20 | 112722 | 50975 | 61747 |  | 28-Sep-20 | 207207 | 50717 | 156490 |

|  |  | ITALY |  |  |  | ITALY |  |  |
| --- | --- | --- | --- | --- | --- | --- | --- | --- |
| Date | Affected | Infected | Removed |  | Date | Affected | Infected | Removed |
| 30-Jun-20 | 240436 | 16496 | 223940 |  | 14-Aug-20 | 249354 | 8695 | 240659 |
| 01-Jul-20 | 240578 | 15563 | 225015 |  | 15-Aug-20 | 249543 | 8605 | 240937 |
| 02-Jul-20 | 240760 | 15255 | 225505 |  | 16-Aug-20 | 249730 | 8517 | 241213 |
| 03-Jul-20 | 240961 | 15060 | 225901 |  | 17-Aug-20 | 249916 | 8431 | 241485 |
| 04-Jul-20 | 241184 | 14884 | 226300 |  | 18-Aug-20 | 250101 | 8346 | 241755 |
| 05-Jul-20 | 241419 | 14621 | 226798 |  | 19-Aug-20 | 250284 | 8263 | 242022 |
| 06-Jul-20 | 241611 | 14642 | 226969 |  | 20-Aug-20 | 250466 | 8181 | 242286 |
| 07-Jul-20 | 241819 | 14709 | 227110 |  | 21-Aug-20 | 250647 | 8100 | 242547 |
| 08-Jul-20 | 241956 | 14242 | 227714 |  | 22-Aug-20 | 250827 | 8020 | 242807 |
| 09-Jul-20 | 242149 | 13595 | 228554 |  | 23-Aug-20 | 251005 | 7942 | 243063 |
| 10-Jul-20 | 242363 | 13459 | 228904 |  | 24-Aug-20 | 251182 | 7865 | 243317 |
| 11-Jul-20 | 242639 | 13428 | 229211 |  | 25-Aug-20 | 251358 | 7789 | 243569 |
| 12-Jul-20 | 242827 | 13303 | 229524 |  | 26-Aug-20 | 251532 | 7714 | 243818 |
| 13-Jul-20 | 243061 | 13179 | 229882 |  | 27-Aug-20 | 251705 | 7640 | 244065 |
| 14-Jul-20 | 243230 | 13157 | 230073 |  | 28-Aug-20 | 251876 | 7567 | 244309 |
| 15-Jul-20 | 243344 | 12919 | 230425 |  | 29-Aug-20 | 252046 | 7495 | 244551 |
| 16-Jul-20 | 243506 | 12493 | 231013 |  | 30-Aug-20 | 252215 | 7424 | 244791 |
| 17-Jul-20 | 243712 | 12305 | 231408 |  | 31-Aug-20 | 252382 | 7354 | 245029 |
| 18-Jul-20 | 243921 | 12120 | 231800 |  | 01-Sep-20 | 252548 | 7284 | 245264 |
| 19-Jul-20 | 244129 | 11942 | 232187 |  | 02-Sep-20 | 252713 | 7216 | 245497 |
| 20-Jul-20 | 244336 | 11768 | 232568 |  | 03-Sep-20 | 252876 | 7148 | 245728 |
| 21-Jul-20 | 244544 | 11600 | 232944 |  | 04-Sep-20 | 253037 | 7081 | 245956 |
| 22-Jul-20 | 244751 | 11437 | 233314 |  | 05-Sep-20 | 253198 | 7015 | 246183 |
| 23-Jul-20 | 244958 | 11279 | 233679 |  | 06-Sep-20 | 253356 | 6949 | 246407 |
| 24-Jul-20 | 245165 | 11126 | 234039 |  | 07-Sep-20 | 253514 | 6884 | 246630 |
| 25-Jul-20 | 245372 | 10978 | 234394 |  | 08-Sep-20 | 253670 | 6820 | 246850 |
| 26-Jul-20 | 245578 | 10833 | 234745 |  | 09-Sep-20 | 253825 | 6756 | 247068 |
| 27-Jul-20 | 245784 | 10693 | 235091 |  | 10-Sep-20 | 253978 | 6693 | 247284 |
| 28-Jul-20 | 245989 | 10557 | 235432 |  | 11-Sep-20 | 254130 | 6631 | 247498 |
| 29-Jul-20 | 246194 | 10424 | 235770 |  | 12-Sep-20 | 254280 | 6569 | 247711 |
| 30-Jul-20 | 246398 | 10296 | 236103 |  | 13-Sep-20 | 254429 | 6508 | 247921 |
| 31-Jul-20 | 246602 | 10170 | 236432 |  | 14-Sep-20 | 254577 | 6448 | 248129 |
| 01-Aug-20 | 246804 | 10048 | 236757 |  | 15-Sep-20 | 254723 | 6387 | 248335 |
| 02-Aug-20 | 247006 | 9929 | 237078 |  | 16-Sep-20 | 254868 | 6328 | 248540 |
| 03-Aug-20 | 247207 | 9812 | 237395 |  | 17-Sep-20 | 255011 | 6269 | 248742 |
| 04-Aug-20 | 247408 | 9699 | 237709 |  | 18-Sep-20 | 255153 | 6210 | 248943 |
| 05-Aug-20 | 247607 | 9588 | 238019 |  | 19-Sep-20 | 255294 | 6152 | 249141 |
| 06-Aug-20 | 247805 | 9480 | 238325 |  | 20-Sep-20 | 255433 | 6095 | 249338 |
| 07-Aug-20 | 248003 | 9375 | 238628 |  | 21-Sep-20 | 255571 | 6038 | 249533 |
| 08-Aug-20 | 248199 | 9271 | 238928 |  | 22-Sep-20 | 255707 | 5981 | 249726 |
| 09-Aug-20 | 248394 | 9170 | 239224 |  | 23-Sep-20 | 255843 | 5925 | 249918 |
| 10-Aug-20 | 248589 | 9071 | 239517 |  | 24-Sep-20 | 255976 | 5869 | 250107 |
| 11-Aug-20 | 248782 | 8974 | 239807 |  | 25-Sep-20 | 256109 | 5814 | 250295 |
| 12-Aug-20 | 248974 | 8879 | 240094 |  | 26-Sep-20 | 256240 | 5759 | 250481 |
| 13-Aug-20 | 249165 | 8786 | 240378 |  | 27-Sep-20 | 256370 | 5705 | 250665 |
| 14-Aug-20 | 249354 | 8695 | 240659 |  | 28-Sep-20 | 256498 | 5651 | 250848 |

|  | USA |  |  |  |  | USA |  |  |
| --- | --- | --- | --- | --- | --- | --- | --- | --- |
| Date | Affected | Infected | Removed |  | Date | Affected | Infected | Removed |
| 30-Jun-20 | 2590668 | 1758754 | 831914 |  | 14-Aug-20 | 5629506 | 3455419 | 2174087 |
| 01-Jul-20 | 2636414 | 1788351 | 848063 |  | 15-Aug-20 | 5714005 | 3499104 | 2214900 |
| 02-Jul-20 | 2687588 | 1829489 | 858099 |  | 16-Aug-20 | 5798406 | 3542182 | 2256223 |
| 03-Jul-20 | 2742049 | 1831276 | 910773 |  | 17-Aug-20 | 5882639 | 3584591 | 2298048 |
| 04-Jul-20 | 2795361 | 1875515 | 919846 |  | 18-Aug-20 | 5966637 | 3626271 | 2340366 |
| 05-Jul-20 | 2841241 | 1817227 | 1024014 |  | 19-Aug-20 | 6050334 | 3667165 | 2383169 |
| 06-Jul-20 | 2891124 | 1854401 | 1036723 |  | 20-Aug-20 | 6133669 | 3707222 | 2426447 |
| 07-Jul-20 | 2936077 | 1881644 | 1054433 |  | 21-Aug-20 | 6216582 | 3746392 | 2470190 |
| 08-Jul-20 | 2996098 | 1928142 | 1067956 |  | 22-Aug-20 | 6299016 | 3784629 | 2514387 |
| 09-Jul-20 | 3054699 | 1968937 | 1085762 |  | 23-Aug-20 | 6380917 | 3821890 | 2559027 |
| 10-Jul-20 | 3117946 | 2015545 | 1102401 |  | 24-Aug-20 | 6462235 | 3858136 | 2604099 |
| 11-Jul-20 | 3184573 | 2067296 | 1117277 |  | 25-Aug-20 | 6542921 | 3893331 | 2649590 |
| 12-Jul-20 | 3245925 | 2115572 | 1130353 |  | 26-Aug-20 | 6622930 | 3927442 | 2695488 |
| 13-Jul-20 | 3304942 | 2163411 | 1141531 |  | 27-Aug-20 | 6702219 | 3960439 | 2741780 |
| 14-Jul-20 | 3364157 | 2196652 | 1167505 |  | 28-Aug-20 | 6780749 | 3992297 | 2788452 |
| 15-Jul-20 | 3431574 | 2246010 | 1185564 |  | 29-Aug-20 | 6858483 | 4022992 | 2835491 |
| 16-Jul-20 | 3497847 | 2284558 | 1213289 |  | 30-Aug-20 | 6935388 | 4052503 | 2882884 |
| 17-Jul-20 | 3550655 | 2310668 | 1239986 |  | 31-Aug-20 | 7011431 | 4080815 | 2930616 |
| 18-Jul-20 | 3605749 | 2338479 | 1267270 |  | 01-Sep-20 | 7086585 | 4107911 | 2978674 |
| 19-Jul-20 | 3662614 | 2367725 | 1294889 |  | 02-Sep-20 | 7160822 | 4133780 | 3027042 |
| 20-Jul-20 | 3721274 | 2398415 | 1322859 |  | 03-Sep-20 | 7234121 | 4158414 | 3075707 |
| 21-Jul-20 | 3781757 | 2430559 | 1351198 |  | 04-Sep-20 | 7306459 | 4181805 | 3124653 |
| 22-Jul-20 | 3844091 | 2464168 | 1379922 |  | 05-Sep-20 | 7377818 | 4203950 | 3173867 |
| 23-Jul-20 | 3908304 | 2499253 | 1409050 |  | 06-Sep-20 | 7448181 | 4224847 | 3223334 |
| 24-Jul-20 | 3974318 | 2535720 | 1438598 |  | 07-Sep-20 | 7517534 | 4244496 | 3273038 |
| 25-Jul-20 | 4042055 | 2573474 | 1468582 |  | 08-Sep-20 | 7585865 | 4262899 | 3322966 |
| 26-Jul-20 | 4111435 | 2612420 | 1499015 |  | 09-Sep-20 | 7653163 | 4280061 | 3373102 |
| 27-Jul-20 | 4182376 | 2652464 | 1529912 |  | 10-Sep-20 | 7719421 | 4295988 | 3423433 |
| 28-Jul-20 | 4254796 | 2693511 | 1561285 |  | 11-Sep-20 | 7784631 | 4310688 | 3473943 |
| 29-Jul-20 | 4328611 | 2735466 | 1593145 |  | 12-Sep-20 | 7848789 | 4324170 | 3524619 |
| 30-Jul-20 | 4403738 | 2778236 | 1625503 |  | 13-Sep-20 | 7911892 | 4336446 | 3575446 |
| 31-Jul-20 | 4480091 | 2821724 | 1658366 |  | 14-Sep-20 | 7973938 | 4347528 | 3626410 |
| 01-Aug-20 | 4557583 | 2865839 | 1691744 |  | 15-Sep-20 | 8034926 | 4357429 | 3677497 |
| 02-Aug-20 | 4636129 | 2910486 | 1725643 |  | 16-Sep-20 | 8094859 | 4366165 | 3728694 |
| 03-Aug-20 | 4715640 | 2955573 | 1760068 |  | 17-Sep-20 | 8153737 | 4373751 | 3779986 |
| 04-Aug-20 | 4796032 | 3001007 | 1795025 |  | 18-Sep-20 | 8211566 | 4380205 | 3831361 |
| 05-Aug-20 | 4877216 | 3046700 | 1830516 |  | 19-Sep-20 | 8268349 | 4385544 | 3882805 |
| 06-Aug-20 | 4959105 | 3092561 | 1866545 |  | 20-Sep-20 | 8324093 | 4389787 | 3934305 |
| 07-Aug-20 | 5041615 | 3138502 | 1903112 |  | 21-Sep-20 | 8378803 | 4392954 | 3985849 |
| 08-Aug-20 | 5124658 | 3184439 | 1940219 |  | 22-Sep-20 | 8432487 | 4395064 | 4037424 |
| 09-Aug-20 | 5208152 | 3230288 | 1977865 |  | 23-Sep-20 | 8485155 | 4396138 | 4089017 |
| 10-Aug-20 | 5292012 | 3275965 | 2016047 |  | 24-Sep-20 | 8536814 | 4396197 | 4140617 |
| 11-Aug-20 | 5376157 | 3321393 | 2054765 |  | 25-Sep-20 | 8587474 | 4395262 | 4192212 |
| 12-Aug-20 | 5460507 | 3366493 | 2094013 |  | 26-Sep-20 | 8637146 | 4393356 | 4243790 |
| 13-Aug-20 | 5544982 | 3411193 | 2133789 |  | 27-Sep-20 | 8685841 | 4390501 | 4295340 |
| 14-Aug-20 | 5629506 | 3455419 | 2174087 |  | 28-Sep-20 | 8733570 | 4386719 | 4346852 |

## Acknowledgements

Thanks are due to Nitay Alon for joint work and to Eytan Ruppin and Laura Sacerdote for helpful suggestions.

The data analyzed in this work is taken from the COVID-19 Data Repository by the Center for Systems Science and Engineering (CSSE) at Johns Hopkins University.

